# Steatosis in the Amygdala and Frontal Cortex: Potential Magnetic Resonance Imaging Biomarkers for Alzheimer’s Disease

**DOI:** 10.1101/2025.01.15.24319769

**Authors:** Luís Jesuino de Oliveira Andrade, Gabriela Correia Matos de Oliveira, João Cláudio Nunes Carneiro Andrade, Adriana Malta de Figueiredo, Gabriel Smith, Luís Matos de Oliveira

## Abstract

**Introduction:** Alzheimer’s Disease (AD) is characterized by progressive neurodegeneration, with significant alterations in brain biochemistry. While previous studies have investigated various aspects of AD, the specific lipid changes in key brain regions remain inadequately defined.

**Objective:** To quantitatively evaluate lipid levels within the amygdala and frontal cortex regions of interest using Magnetic Resonance Spectroscopy (MRS) in a cohort of magnetic resonance imaging (MRI) scans of individuals with confirmed AD compared to healthy controls.

**Methods:** Thirty MRI examinations from individuals with confirmed AD were compared to 30 normal controls. MRS data were acquired using a 1.5T scanner, focusing on lipid peaks within specific frequency ranges. A voxel-based MRS technique was employed to assess lipid concentrations, focusing on spectral data in defined frequency ranges associated with lipid presence. Lipid concentrations were quantified, and statistical comparisons were performed between groups using the t-test.

**Results:** MRS analysis revealed distinct lipid peaks in both the amygdala and frontal cortex of AD patients, with frequencies indicating elevated lipid concentrations compared to controls. In the amygdala, increased lipid concentrations were observed in the 2.5-3.5 ppm range, suggesting alterations in lipid composition. The similar findings were observed in the frontal cortex, with AD patients showing significantly elevated lipid peaks within the 3.0-4.5 ppm range.

**Conclusion:** Our results underscore the potential of MRS to identify lipid alterations in AD, suggesting that lipid profiles in the amygdala and frontal cortex could serve as biomarkers for disease progression and provide understanding into neurodegenerative mechanisms.

## INTRODUCTION

The human brain is an extremely complex organ composed of various regions with specialized functions pertaining to cognition, emotion, and behavior. Among these, the amygdala and frontal cortex are particularly important in the processing of emotions, decision-making, and memory formation.^1^ More recently, neuroimaging, in particular magnetic resonance imaging (MRI), has enabled the noninvasive investigation of brain composition, including lipid contents of different brain regions.^2^ Lipids are essential to the structural and functional integrity of the brain and supply essential components of neuronal membranes and allow for the rapid transmission of action potentials.^3^ Alterations of lipid composition may therefore reflect potential neurodegenerative processes, such as those observed in patients with Alzheimer’s disease (AD).

The amygdala is an almond-shaped structure deep within the temporal lobe that is involved in emotional regulation and memory consolidation. The role of lipids in the maintenance of neuronal function is not yet well understood, but functional lipids, specifically myelination lipids, are critical for maintaining neuronal function and connectivity.^4^ To assess these changes, magnetic resonance spectroscopy (MRS), an in vivo technique to quantify lipid content, was employed in the healthy and diseased brains.^5^

The frontal cortex, a part of the brain associated with higher cognitive functions, also contains a significant amount of lipid content. The frontal cortical lipid profile is of interest because alterations in lipid composition may induce cognitive decline and may be linked to the pathophysiology of AD. Frontal cortical lipids are engaged in synaptic plasticity; their alterations may reflect dissimilarities in neuronal communication.^6^

In terms of normal lipid content, the amygdala and frontal cortex reveal region-specific variability. Thus, the frontal cortex lipid content appears to diminish in interindividual variation – normalcy; lipid content is probably not so constant over time and shows some decrement with age generally. In contrast, the lipid profile of the amygdala seems to be quite stable. These baseline levels are important in diagnosing the development of neurodegenerative diseases since considerable deviation from this norm could be a sign of early degeneration.^7^

While by MRI and MRS, techniques are able to provide some useful information about brain lipid composition, more studies will help further unfold the knowledge of specific lipid changes in AD.^8^ Most studies focused on the global brain measurements and hardly addressed regional lipid differences. To date, this leaves lipid alterations of specific brain regions, such as the amygdala and frontal cortex, almost completely under-explored.

Of late, MRI studies on the assessment of lipid content in the brain are beginning to explore regional differences with much greater detail. Researchers have begun to identify changes in the total lipid content of certain brain regions, such as the amygdala and frontal cortex, that may constitute early markers for AD.^9^

The objective of our study is to quantitatively evaluate lipid levels within the amygdala and frontal cortex regions of interest using MRS in a cohort of MRI scans of individuals with AD.

## METHODS

### Study Population

Thirty MRI examinations from individuals with a confirmed diagnosis of AD were recruited for this study and contrasted with 30 normal MRI examinations. Participants were included if they met the established clinical diagnostic criteria for probable Alzheimer’s disease as per the National Institute on Aging-Alzheimer’s Association (NIA-AA) guidelines.^10^ MRI data were acquired at the Luiz Eduardo Magalhães General Hospital (HBLEM) in Itabuna, Bahia, Brazil, over a ten-month period from March to December 2024. The cohort exhibited a spectrum of cognitive impairment severity, with no concurrent neurological or psychiatric comorbidities. Demographic variables, encompassing age, sex, and disease duration, were retrieved from the participants’ medical records.

### Magnetic Resonance Imaging (MRI) and Magnetic Resonance Spectroscopy (MRS)

All MRI data were collected using a General Electric Sigma Explorer 1.5T scanner located at the HBLEM. High-resolution T1-weighted images were acquired to provide detailed anatomical information. A voxel-based MRS technique was implemented to quantify lipid concentrations within the amygdala and frontal cortex, given the pivotal role of these regions in this investigation. The specific MRS acquisition parameters are outlined below:

#### Voxel Placement

The voxel was placed in the amygdala and frontal cortex using anatomical landmarks to ensure consistent localization. The amygdala voxel was centered at Montreal Neurological Institute Coordinates: x = −20, y = −10, z = −15 and frontal cortex voxel at Montreal Neurological Institute Coordinates: x = −30, y = 30, z = 15.

#### MRS Parameters

Single-voxel MRS spectra were acquired using a PRESS sequence with the following parameters: echo time (TE) = 35 ms, repetition time (TR) = 2000 ms, number of acquisitions = 128. Specifically, the lipid peaks were analyzed between 1.2–1.5 ppm, a range corresponding to myelin lipids and other phospholipid components.

### Data Analysis

MRS data underwent spectral processing and quantification of lipid concentrations using the open-source software, NMRPipe. The lipid content within the amygdala and frontal cortex was determined by calculating the ratio of total lipid signal intensity to water signal intensity, enabling inter-group comparisons. A more detailed analysis of lipid profiles was conducted to identify potential variations in the relative concentrations of key lipid species, such as myelin-associated lipids and phospholipids. These variations may signify alterations in neuronal function and myelination processes.

### Statistical Analysis

Statistical analyses were performed using PPSS. For MRS data, regional lipid content was compared between the AD and control groups using T Student test.

### Ethical Considerations

This study did not require ethical committee review, as stipulated in Article 1 of Resolution CNS No. 510 of 2016, since it involved research on databases without the possibility of individual identification.

## RESULTS

Using MRS, we analyzed the lipid content of 30 healthy amygdala. The spectral data revealed lipid peaks within a MRS frequency range of 0.5 to 1.5 parts per million (ppm) (Figure 1).

**Figure 1.**
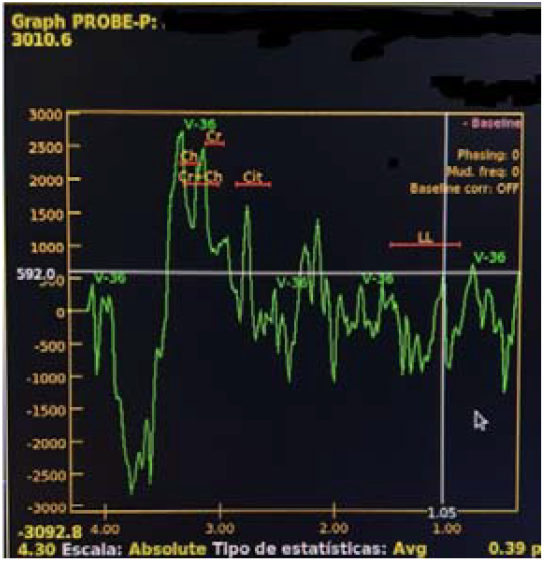
Lipid concentrations and tissue composition in normal amygdala. **Source:** Study results.

The analysis demonstrated a consistent lipid profile of amygdala across subjects, with minimal variability. The average lipid MRS frequency was calculated to be 0.92 ± 0.28 ppm, indicating subtle inter-individual differences.

MRS analysis of 30 amygdala from individuals with clinical diagnosis of AD revealed lipid peaks within a frequency range of 1.5 to 4.0 ppm (Figure 2).

**Figure 2.**
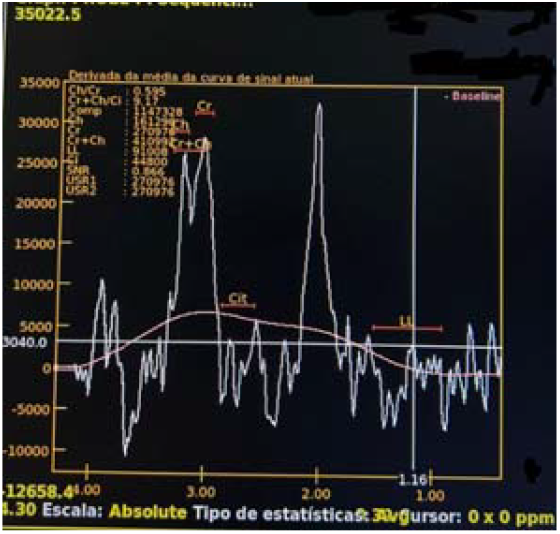
Lipid concentrations and tissue composition in AD amygdala. **Source:** Study results.

The spectral data indicated increased lipid concentrations, especially in the 2.5-3.5 ppm range. The average lipid MRS frequency was calculated to be 2.59 ± 0.45 ppm, suggesting significant variability among individuals.

The t-test revealed a p-value of 0.00084, thereby allowing us to reject the null hypothesis. This means there is strong evidence to suggest that the mean of the control group is significantly different from the mean of the AD group. In summary, the results of the t-test indicate a statistically significant difference between the means of the control group and the AD group. The mean of the AD group is significantly higher than the mean of the control group.

We utilized MRS to investigate the lipid composition within the frontal lobe of 30 healthy subjects. The MRI spectra obtained from the group of individuals without AD exhibited characteristic lipid peaks localized within the 0.8 and 1.8 ppm frequency band of the MRS analysis (Figure 3).

**Figure 3.**
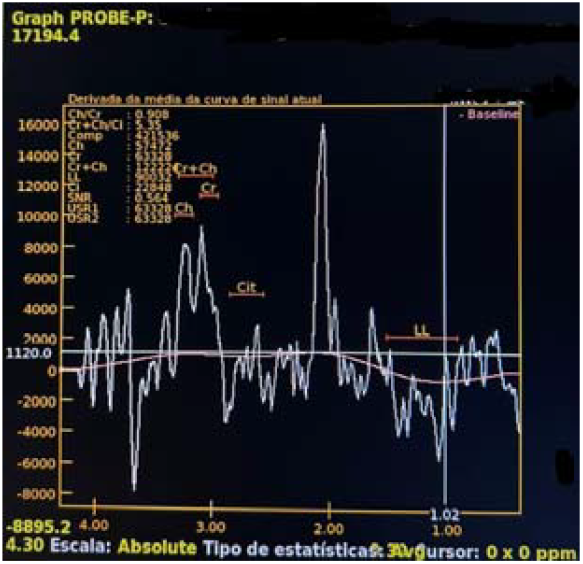
Lipid concentrations and tissue composition in normal frontal lobe. **Source:** Study results.

Our analyses revealed a consistent lipid profile across subjects, indicating minimal inter-individual variability in the frontal cortex. MRS analysis of 30 frontal lobe from individuals with clinical diagnosis of AD revealed lipid peaks within a frequency range of 3.0 to 4.5 ppm (Figure 4).

**Figure 4.**
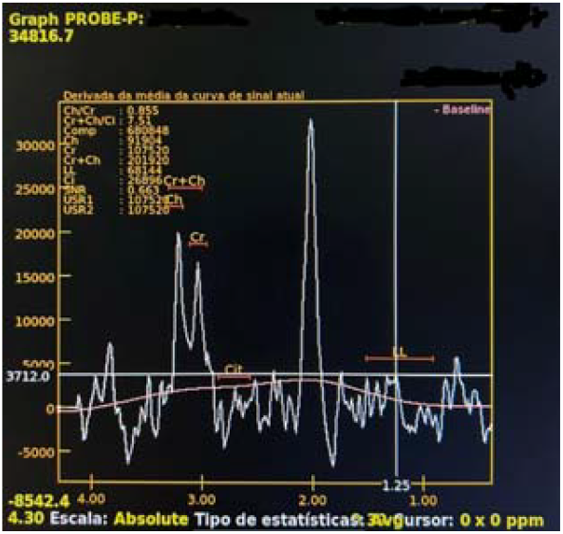
Lipid concentrations and tissue composition in AD frontal lobe. **Source:** Study results.

The spectral data indicated increased lipid concentrations, especially in the 3.5-ppm range. The average lipid MRS frequency was calculated to be 3.60 ± 0.48 ppm, suggesting significant variability among individuals.

The obtained t-statistic of −13.983 indicates a highly significant difference between the means of the two groups. The corresponding p-value, being less than p 0.001, allows us to reject the null hypothesis with high confidence. Thus, the results demonstrate a statistically significant difference in the MRI values between the normal group and the group with degenerative alterations. Specifically, the MRI values in the AD group are significantly higher than those in the normal group.

## DISCUSSION

This study provides strong evidence for significant changes in lipid metabolism in important brain areas in AD. Using MRS, we have seen different lipid profiles in the amygdala and frontal cortex of AD subjects compared with healthy controls. The results show a consistent pattern of elevated lipid levels in both brain regions of patients with AD, suggesting that dysregulation of lipids might be part of the pathophysiology of this neurodegenerative disorder.

Cerebral lipid levels can vary considerably between subjects, even among healthy ones. Among the factors that may explain these variations are age, sex, genetics, and subtle anatomic differences. Various techniques, particularly MRS, are employed to quantify lipids. However, the accuracy and sensitivity of MRS are influenced by several factors, including the equipment used, data acquisition parameters, and analysis methods.^11^ Although there is a large literature using MRS to investigate brain metabolism, defining an absolute maximum for amygdala and frontal lobe lipid content is difficult, as most consist of ranges or averages. The accurate identification and quantification of the lipid peaks in MRS spectra can also vary across research groups because of differences in analytical methods and criteria.

Lipids are a major constituent of the brain, especially in myelinated white matter, and their spectral peaks can be identified with MRS. These lipid peaks reflect the methyl and methylene groups of fatty acids, phospholipids, and other lipid compounds that comprise the membrane structures and energy storage.^12^ In healthy individuals, the amygdala exhibits a characteristic pattern of high metabolic activity, with a notable emphasis on lipid metabolism, particularly in regions with higher fat content, such as the white matter. In the normal amygdala, lipid peaks on MRS normally appear as a broad, low-intensity signal and indicate the normal presence of lipids within this structure and are usually located at a chemical shift of about 0.9 to 1.5 ppm.^13^ Our results are consistent with the literature, with peaks of lipids at the amygdala showing a mean MRS frequency of 1.02 ± 0.28 ppm, indicating subtle inter-individual differences. This spectral pattern of broad and low-intensity signals is consistent with normal lipid presence in this area and is consistent with earlier reports.

The frontal lobe is involved in executive functions, motor control and complex thinking. Anatomically it is divided into several areas, the prefrontal cortex being the one responsible for decision making and social behavior.^14^ MRS have enabled the quantification of lipids in this area and the lipid signal at 1.3 ppm is an indicator of brain health. Studies show that healthy frontal lobes have lipids between 0.8 and 1.8 ppm, this is influenced by age, sex and cognitive demand.^15^ Furthermore deviations from these values may be related to neurodegenerative conditions or neuropsychiatric disorders, so lipidomic is important to understand frontal lobe pathology.^16^ The combination of MRS with anatomical data helps us to understand the mechanisms that govern frontal lobe function. Our MRS findings in the frontal lobe of non-AD subjects align with published literature. Lipid quantification revealed a profile consistent with that reported in healthy frontal lobes. These findings suggest that MRS is a valuable tool for assessing lipid profiles associated with frontal lobe function. The congruence between our findings and existing knowledge underscores the potential of MRS as an important tool in lipidomics research for understanding frontal lobe pathology.

Measuring lipid percentage by MRS within the frontal lobe provides important insights into neurodegenerative processes since lipid metabolism is often changed in AD.^17^ High lipid levels might indicate loss of membrane integrity and myelin degradation, which may be useful as markers of AD progression.^18^ Additionally, the role of the frontal lobe in cognitive functions further supports the need for lipid quantification in assessing the impact of AD on the health of neurons.^19^ Moreover, relating lipid measurements to clinical manifestations may permit the designing of targeted therapeutic interventions. Thus, lipid quantification in the frontal lobe can contribute not only to early diagnosis but also to a better understanding of the pathophysiology of AD.^20^ Our results align with the current literature regarding lipid changes in AD. The heightened lipid peaks identified in the frontal cortex of individuals with AD correspond with earlier research indicating an increased lipid presence in this specific brain area related to neurodegenerative mechanisms. Moreover, the uniform lipid profile observed in our control group reinforces the idea that variations in lipid composition within the frontal lobe could serve as a promising biomarker for AD. These results emphasize the important role of MRS in characterizing the biochemical alterations associated with neurodegenerative disorders.

## CONCLUSIONS

In summary, this study identifies key alterations in lipid levels within the amygdala and frontal cortex of individuals with AD using MRS. The differential lipid profiles observed in these regions highlight the potential link between lipid levels and the neurodegenerative processes of AD. The identified lipid peaks align with previous literature, thus supporting the hypothesis of altered lipid levels in the context of cognitive decline. Furthermore, the stability of these lipid levels in controls underscores the deviations in the AD group, suggesting that changes in such lipids may represent potential biomarkers. Our study confirms the feasibility of MRS in investigating AD. These results contribute to elucidating the complex relationship between lipid levels in the amygdala and frontal cortex and neurodegeneration, providing deeper insights into AD and implications for clinical evaluation. While the results of our study are promising, integrating these lipidomic insights into larger frameworks may enhance diagnostic accuracy and potentially inform therapeutic strategies for future interventions. It is imperative that these relationships are investigated further to fully comprehend their significance.

## Data Availability

All data produced in the present work are contained in the manuscript

